# ‘It’s only a flesh wound’ – understanding the safety culture in equine, production animal and mixed veterinary practices

**DOI:** 10.1101/2025.01.06.25320051

**Authors:** John S.P. Tulloch, Imogen Schofield, Rebecca Jackson, Martin Whiting

## Abstract

The veterinary industry has some of the highest rates of non-fatal work-related injuries, yet safety culture remains unexplored. Utilising a survey, this study aimed to describe the prevalence of work-related injuries in equine, production animal, and mixed veterinary practices, and to understand the behaviours of injured persons.

There were 144 respondents. Over 90% of clinicians experienced injury during their careers, versus a third of non-clinical staff. Injuries to equine veterinarians were predominately kicks to the leg or head, and usually involved the examination of a horse’s distal limb, most were not wearing hard hats. Production animal veterinarians’ injuries included crushed hands and feet, and kicks to legs. Injuries often lead to hospital attendance (>25% equine, >40% production animal), yet few took time off work.

Veterinarians avoided taking time off work after injuries, reasons included; minimisation of injury severity, not wanting to ‘let the team down’, and feelings of guilt. Many planned behaviour change post-injury, including using protective headwear, increasing awareness of surroundings, and using better restraint. Most injuries went unreported due to lack of awareness, acceptance of injury risk, time constraints, and believing reporting would have no impact.

This study exposes a culture in large animal veterinary workplaces that normalises injuries and undervalues safety. The lack of protective measures and the tendency to continue working despite serious injuries, highlight a need for urgent cultural and systemic change. Improved safety practices, leadership commitment, and comprehensive training are essential to address this deep-seated issue and fostering a safer work environment.

## Introduction

The veterinary industry has some of the highest rates of non-fatal occupational injuries. In the USA, in 2022, it lies second out of all industries (10.2 injuries per 100 fulltime equivalent (FTE) workers) behind skiing facilities (12.0 per 100 FTE workers) and is more than four times higher than the national average (Bureau of Labor Statistics, 2023). In the UK around two-thirds of veterinary surgeons are injured annually (Mills, 2019). The main hazard associated with veterinary surgeon injuries are the animals being treated (Gabel and Gerberich, 2002; Johnson and Fritschi, 2024; Tulloch et al., 2023). For equine and production animal veterinarians the main hazards are horses and cattle, which can cause fatal and life changing injuries. Despite the high-risk nature of working with these species, research has primarily focused on the general equestrian or agricultural sector with little exploring the veterinary industry.

When equine-related injuries are discussed, predominately the focus is on riding related injuries, however a significant proportion occur when unmounted. These will be the type of injuries that veterinarians are subject to. Recent studies show that more than 20% of all horse-related injuries were caused by a horse kick when unmounted, with a further 10% caused by a trampling (Lang et al., 2014; Neville et al., 2024). The main anatomy injured by kicks are a person’s head, thorax or abdomen (Franzén Lindgren et al., 2023; Neville et al., 2024; Rutty and Cheshire, 2017; Savage et al., 2023; Sritharan et al., 2022; Wolyncewicz et al., 2018). The force involved in such a kick can be 1.8 times a horse’s body weight (around 10,000 Newtons), and it is no surprise that half of all such head injuries lead to traumatic brain injuries (Neville et al., 2024; Rutty and Cheshire, 2017). A study found that head injuries are just as likely between mounted and unmounted injuries, (Carmichael et al., 2014) and that helmeted equestrians receive less severe traumatic brain injuries than un-helmeted individuals (Bier et al., 2018). Despite this only a small proportion of individuals wear helmets (<8%) when unmounted (Eckert et al., 2011). Trampling and crush injuries often lead to severe contusions, multiple fractures and trauma to chest and abdominal organs (Neville et al., 2024; Rutty and Cheshire, 2017; Thomas et al., 2006).

On average six people are killed by cattle each year in the UK (Health and Safety Executive, 2023), and 20-22 in the USA (Byard, 2024). These are predominately agricultural workers, but do include members of the public, mainly dog walkers. Injuries sustained are often classified as major traumas, with high levels of hospital admission often with intensive care involvement, with injury severity scores equivalent to high-speed road traffic accidents (Murphy et al., 2010; Rhind et al., 2021; Sheehan et al., 2020). The mechanisms of injury are predominately being trampled, indirect crushes/blunt trauma, and kicks (Ehrhard et al., 2022; Iwai and Yamamoto, 2023; Murphy et al., 2010; Rhind et al., 2021; Savage et al., 2023; Sheehan et al., 2020; Tosswill et al., 2018). There is a high fracture rate (40-46%) mainly to upper limbs and spine, and many head injuries (2-86%), ranging in severity from concussions to facial fractures.

Research has yet to fully explore the context of these injuries as a means to develop preventative practices, nor explored the health and safety culture, within the veterinary profession. A UK study of equine veterinarians found that 33% of their worst injuries resulted in a hospital admission, and 7% reported a loss of consciousness. These were mainly caused by kicks related to distal limb or head examinations (Parkin et al., 2018). An audit of UK veterinary schools identified that 16% of those injured by horses, and 21% by cattle, required a hospital visit (Tulloch et al., 2023). Clinicians injured by horses were performing a clinical examination, sedating an animal or restraining an animal, whilst those injured by cattle were performing a clinical examination, injured by a ‘free animal’, or performing a veterinary procedure (Tulloch et al., 2023).

Emerging evidence has highlighted that how veterinarians perceive workplace injuries is concerning. One in five equine, and more than one in three production animal veterinarians, believe that for something to qualify as an injury it must impact their ability to work, additionally 13% of production animal veterinarians stated an injury needed to require medical treatment (Furtado et al., 2024). Similarly, a large proportion of both types of veterinarians would only report an injury if it impacted their ability to work or that it was severe. Concerningly, a proportion would not report injuries if they attributed themselves as the cause of the injury. Some within the veterinary industry are starting to realise the issues that exist and have called for standardised professional workplace safety guidelines (Potter et al., 2024). But with a paucity of contextual and cultural data, these will be challenging to create.

The aim of this study was to describe the prevalence of work-related injuries in equine, production animal, and mixed veterinary practices, and to understand the decision-making behaviour of injured persons in these settings.

## Materials and Methods

An online cross-sectional survey was designed and distributed to all UK employees of a consolidated group of veterinary practices, who work in equine, production animal, and mixed animal clinical practices. The survey was distributed through weekly email newsletters and was open between 6^th^ December 2022 and 6^th^ March 2023. The survey was piloted with a small group of veterinarians, nurses and health and safety professionals. To enhance recruitment, an incentive of a year’s supply of tea, coffee, and biscuits, for the practices of five randomly selected respondents was provided.

Respondents to the survey were asked about personal demographics, job role details, working hours, and whether they perform clinical work. Respondents were asked whether they had acquired a work-related injury during their career in the veterinary sector. If they had, they were presented with a series of questions about their most recent injury and their most severe injury. These included questions around the context of how the injury was acquired, how long ago the injury occurred, the medical consequences of injury, any resultant behaviour change, whether they took time off work, and whether they reported their injury to their employer. Injury was defined as

> ‘A bodily harm resulting from severe exposure to an external force or substance (mechanical, thermal, electrical, chemical, or radiant) or a submersion. This bodily harm can be unintentional or violence-related.(’CDC, 2024)

We stated that for an injury to be work related it must be:

> ‘resultant or in connection with work. The work activity itself must contribute to the injury’. (Health and Safety Executive, 2024a)

Demographic characteristics of respondents were described. Overall injury prevalence was calculated and stratified by job role. Logistic regression was performed to identify any association between demographic variables and receiving an injury. Variables taken forward for multivariable analysis were selected through substantive knowledge and statistical significance (ie where p<=0.3). Hosmer-Lemeshow tests were performed to assess goodness of fit.

The data was subsequently stratified from the remainder of the analysis into the following groups; non-clinical roles in all practices, clinical roles in equine practices, clinical roles in production animal practices, and clinical roles in mixed practices. The types of injury acquired were described and it was recorded whether they were reportable to the Health and Safety Executive (HSE). In the UK, under the Reporting of Injuries, Diseases and Dangerous Occurrences Regulation 2013 (RIDDOR), an employer legally must report work-related accidents that result in specific injuries to the HSE (Health and Safety Executive, 2013). These injuries include any that leave them incapacitated for more than seven days, fractures other than to digits, loss of consciousness, among others. We performed more focused analysis on animal-related injuries, and the procedures occurring at the point of injury, as we anticipated that these would represent most injuries to clinical staff.

The analysis of questions relating to the context of injury, attitude of injured person, and reporting culture were carried out descriptively. Open text questions were analysed using iterative qualitative content analysis (Krippendorff, 2019; Vears and Gillam, 2022). The responses were read through with the researchers making note of any initial impressions; secondly, the researchers carried out initial “coding”, by reading items individually and labelling important concepts. As more text was incorporated into the analysis, codes were refined, combined, or renamed to more accurately represent the information conveyed by respondents. Eventually, codes could be grouped into overall categories or “themes”, which were counted to aid interpretation and comparison.

Data masking was used to protect personally identifiable information of respondents, this was performed through aggregation of data or redacting of quotes. The study received ethical approval from the University of Liverpool Veterinary Research Ethics Committee (VREC1256).

## Results

One hundred and forty-four individuals consented to take part in the survey. This represents a response rate of 32.3%. Due to the low number of veterinary nurses responding to the survey, they were combined with veterinarians to form the ‘clinical staff’ groups (Table 1). Across all veterinary sectors, 78.0% of veterinarians and veterinary nurses are female, the median age group is 30-39, and 97.0% of individuals identify as being white (Robinson et al., 2019b, 2019a). Demographically the clinical respondents were broadly representative. No information exists about the demographics of non-clinical staff.

**Table 1.** Demographics of respondents to a survey on veterinary work-related injuries.

|  | Non-clinical staff<br>(n=40) | Clinical Equine Staff<br>(n=70) | Clinical Production<br>Animal Staff (n=16) | Clinical Mixed Practice<br>Staff (n=18) |
| --- | --- | --- | --- | --- |
| **Sex (Female)** | 95.0% (83.1-99.4) | 72.9% (60.9-82.8) | 81.3% (54.4-95.6) | 77.8% (52.4-93.4) |
| **Age** |  |  |  |  |
| 20-29 | 37.5% (23.6-53.1) | 20.0% (11.4-31.3) | 37.5% (15.2-64.6) | 27.8% (9.7-53.5) |
| 30-39 | 15.0% (5.7-29.8) | 32.9% (22.1-45.1) | 37.5% (15.2-64.6) | 22.2% (6.4-47.6) |
| 40-49 | 25.0% (12.7-41.2) | 30.0% (19.6-42.1) | 18.8% (4.0-45.7) | 33.3% (13.3-59.0) |
| 50+ | 22.5% (10.8-38.5) | 17.1% (9.2-28.0) | 6.3% (0.2-30.2) | 16.7% (3.6-41.4) |
| **Ethnicity** |  |  |  |  |
| White | 100% | 95.7% (88.0-99.1) | 100% | 100% |
| All other ethnic groups combined | 0 | 4.3% (0.9-12.0) | 0 | 0 |
| **Role** |  |  |  |  |
| Administrator | 65.0% (48.3-79.4) | 1.4% (0.0-7.7) | 0 | 5.6% (0.1-27.3) |
| Animal care assistant | 0 | 12.9% (6.1-23.0) | 0 | 0 |
| Receptionist | 22.5% (10.8-38.5) | 0 | 0 | 0 |
| Technician | 12.5% (4.2-26.8) | 7.1% (2.4-15.9) | 0 | 0 |
| Veterinary Nurse | 0 | 11.4% (5.1-21.3) | 0 | 16.7% (3.6-41.4) |
| Veterinary Surgeon | 0 | 67.1% (54.9-77.9) | 100% | 77.8% (52.4-93.4) |
| **Hours of employment** |  |  |  |  |
| <10 hours | 0 | 1.4% (0.0-7.7) | 0 | 0 |
| 11-20 hours | 5.0% (0.6-16.9) | 4.3% (0.9-12.0) | 0 | 0 |
| 21-30 hours | 25.0% (12.7-41.2) | 14.3% (7.1-24.7) | 12.5% (1.6-38.4) | 11.1% (1.4-34.7) |
| 31-40 hour | 65.0% (48.3-79.4) | 40.0% (28.5-52.4) | 37.5% (15.2-64.6) | 5.6% (0.1-27.3) |
| 41-50 hours | 5.0% (0.6-16.9) | 27.1% (17.2-39.1) | 43.8% (19.8-70.1) | 66.7% (41.0-86.7) |
| 50+hours | 0 | 12.9% (6.1-23.0) | 6.3% (0.2-30.2) | 16.7% (3.6-41.4) |
| **Out of hours** |  |  |  |  |
| No out of hours | 75.0% (58.8-87.3) | 22.9% (13.7-34.5) | 0 | 11.1% (1.4-34.7) |
| <10 hours | 22.5% (10.8-38.5) | 34.3% (23.4-46.6) | 31.3% (11.0-58.7) | 33.3% (13.3-59.0) |
| 11-20 hours | 0 | 27.1% (17.2-39.1) | 37.5% (15.2-64.6) | 38.9% (17.3-64.3) |
| 21-30 hours | 0 | 10.0% (4.1-19.5) | 6.3% (0.2-30.2) | 16.7% (3.6-41.4) |
| 31-40 hour | 0 | 2.9% (0.3-9.9) | 12.5% (1.6-38.4) | 0 |
| 41-50 hours | 0 | 0 | 6.3% (0.2-30.2) | 0 |
| 50+hours | 2.5% (0.1-13.2) | 2.9% (0.3-9.9) | 6.3% (0.2-30.2) | 0 |
| **Clinical Responsibilities** |  |  |  |  |
| <10 hours | NA | 15.8% (8.1-26.4) | 0 | 0 |
| 11-20 hours | NA | 8.6% (3.2-17.7) | 18.8% (4.0-45.7) | 16.7% (3.6-41.4) |
| 21-30 hours | NA | 18.6% (10.3-29.7) | 37.5% (15.2-64.6) | 38.9% (17.3-64.3) |
| 31-40 hour | NA | 24.3% (14.8-36.0) | 18.8% (4.0-45.7) | 27.8% (9.7-53.5) |
| 41-50 hours | NA | 22.9% (13.7-34.5) | 18.8% (4.0-45.7) | 11.1% (1.4-34.7) |
| 50+hours | NA | 10.0% (4.1-19.5) | 6.3% (0.2-30.2) | 5.6% (0.1-27.3) |

More than 90% of all clinical staff had received a work-related injury during their career; 94.3% of equine clinical staff, 93.7% of production animal clinical staff, and 100% of mixed practice clinical staff (Figure 1). In contrast, a third (32.5%) of non-clinical staff had received a work-related injury. Production animal and mixed practice clinical staff reported the highest frequency of work-related injuries throughout their careers.

Univariable analysis to identify which demographic variables were associated with a respondent having ever acquired a veterinary work-related injury identified the following variables as significant; sex, age, role, clinical status, and out of hours status. (Table 2) There was no association found for practice type, and knowledge of their practice’s health and safety protocol. When taking forward variables for multivariable analysis, role was removed as there was strong collinearity between it and clinical status. Subsequently, the only significant age group were 50-59 year olds where the odds of having ever received an injury was 8.1 times higher than 20-29 year olds, when adjusted for clinical status. The odds of respondents who performed a clinical role having acquired an injury was 35.5 times higher than those with non-clinical roles, when adjusted for age, sex and out of hours status.

**Table 2.** Univariable and multivariable logistic regression (adjusted for age, sex, clinical and out of hours status) exploring demographic factors associated with acquiring a veterinary work-related injury in large animal practices.

|  | Univariable analysis |  | Multivariable analysis |  |
| --- | --- | --- | --- | --- |
|  | Odds Ratio (95% CI) | p-value | Odds Ratio (95% CI) | p-value |
| **Sex (Ref: Female)** | 4.53 (1.25-29.2) | 0.048 | 2.6 (0.3-4.6) | 0.40 |
| **Age (Ref: 20-29)** |  |  |  |  |
| 30-39 | 5.25 (1.68-20.16) | 0.008 | 4.7 (0.98-26.4) | 0.06 |
| 40-49 | 2.83 (1.03-8.39) | 0.049 | 3.6 (0.85-17.1) | 0.09 |
| 50-59 | 4.50 (1.07-31.1) | 0.067 | 8.1 (1.1-79.7) | 0.05 |
| 60-69 | 0.60 (0.12-2.89) | 0.51 | 0.65 (0.06-7.0) | 0.72 |
| **Practice Type (Ref: Equine)** |  |  |  |  |
| Production Animal | 0.51 (0.20-1.34) | 0.16 |  |  |
| Mixed | 0.87 (0.30-2.89) | 0.80 |  |  |
| **Role (Ref: Receptionist)** |  |  |  |  |
| Animal Care Assistant | 7.0 (0.97-72.6) | 0.07 |  |  |
| Administrator | 0.8 (0.16-4.51) | 0.79 |  |  |
| Veterinary Nurse | 20.0 (2.26-470.67) | 0.018 |  |  |
| Veterinary Surgeon | 75.0 (12.21-711.92) | <0.0001 |  |  |
| Veterinary Technician | 18.0 (2.01-425.66) | 0.023 |  |  |
| Clinical Status<br>(Ref: Non-clinical) | 41.1 (14.5-139.5) | <0.0001 | 35.5 (10.2-157.9) | <0.001 |
| Out of Hours Status<br>(Ref: None) | 6.29 (2.73-15.4) | <0.0001 | 1.3 (0.3-4.6) | 0.72 |
| Knowledge of Health<br>and Safety Protocol<br>(Ref: N) | 0.42 (0.16-1.00) | 0.06 |  |  |

The last 12 months annual work-related injury prevalence for non-clinical staff was 17.5% (95%CI 7.3-32.8), for equine clinical staff it was 48.6% (95%CI 36.4-60.8), for production animal clinical staff 62.5% (95%CI 35.4-84.8), and for mixed practice clinical staff 66.7% (95%CI 41.0-86.7).

Non-clinical staff were predominately injured within the practice (76.9% of recent injuries, 75.0% of severe injuries), whilst clinical staff were mainly injured outside of the practice (61.5% recent, 59% severe) (Table 3).

**Table 3.**
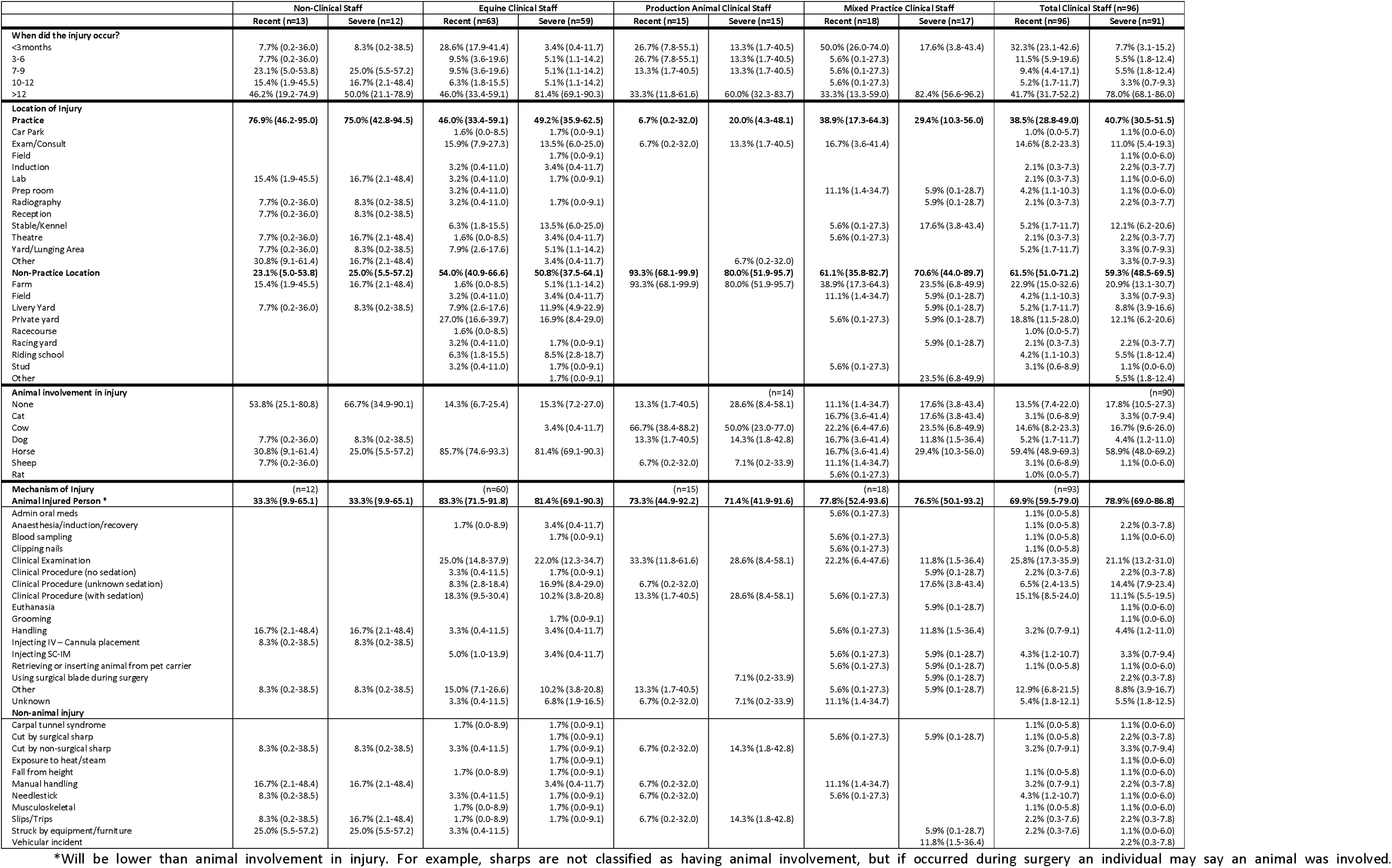
Context of veterinary work-related injuries.

Around 70% of all recent injuries, and almost 80% of severe, acquired by clinical staff had animal involvement, whilst only 33.3% of non-clinical staff did. The main mechanism of injury to non-clinical staff was being ‘struck by equipment or furniture’. Whilst for clinical staff, 49.6% of recent, and 48.8% of severe, were related to a clinical procedure or clinical exam.

The most prevalent animal-related injuries occurring to equine clinical staff were; kicks to the leg (30% recent, 20% severe), kicks to the head (12% recent, 16% severe), a crushed foot (8% recent), a rear landing on a foot (8% severe) (S1). An injury to the head occurred in 16% of recent and 22% of severe injuries. Sixteen of recent, and 26% severe horse related injuries resulted in a fracture.

The most prevalent recent animal-related injuries occurring to production animal clinical staff were; a crushed hand (25% recent), kicks to the leg (16.7% recent), a crushed foot (16.7% recent), and a bite to the hand (16.7% recent). There was no predominant injury mechanism in severe injuries to production animal clinical staff, though 12.5% were to the head. Seventeen percent of recent, and 25.0% of severe, cattle related injuries resulted in a fracture.

The most prevalent animal-related injuries occurring to mixed practice clinical staff were; a bite to the hand (16.3% recent, 22.2% severe), kicks to the leg (18.8% recent, 22.2% severe), a bite to the arm (6.3% recent, 33.3% severe), and a kick to the head (12.5% recent). Over twenty percent (22.2%) of severe injuries resulted in a fracture.

For equine clinical staff, when it was possible to extract the exact procedure involved (recent n=30, severe n=26), 43.3% and 34.6% respectively involved examination of the distal limb horse. The top procedures involved in recent injuries included lifting a hoof for examination (16.7%, 95% CI 5.6-34.7), distal limb nerve blocks (10.0%, 95% 2.1-26.5), ultrasound scanning of the distal limb (10.0%, 95% 2.1-26.5), and endoscopy (10.0%, 95% 2.1-26.5). In severe injuries they were distal limb nerve blocks (19.2%, 95%CI 6.6-39.4) and rectal examination (11.6%, 95% CI 2.4-30.2). When recently injured by a horse 32.1% (95% CI 19.9-46.3) of equine clinical staff reported wearing personal protective equipment (PPE), this was 28.6% (95% CI 17.3-42.2) in severe injuries. In recent injuries, 63.2% wore a helmet, 15.8% wore a body protector, the remainder wore chaps, gloves, or steel toe capped boots. In severe injuries, 58.3% wore a helmet, 25.0% wore steel toe capped boots, and 16.7% wore gloves.

Seven production animal clinical staff could provide details of their most recent injuries, two further could provide details in their most severe injuries. Of the most recent injuries, only two procedures were mentioned more than once; oral examination of cattle (28.6%, 95%CI 3.7-71.0), and castration of cattle (28.6%, 95%CI 3.7-71.0). For the severe injuries they were; caesarean sections of cattle (22.2%, 95%CI 2.8-60.0), and clinical examination of a farm dog (22.2%, 95%CI 2.8-60.0). For mixed practice clinical staff, in both recent and severe injuries, each procedure involved a different species so meaningful conclusions could not be drawn.

Regarding their most recent injury, no non-clinical staff nor mixed practice clinical staff acquired a work-related injury that was reportable to the HSE through RIDDOR as a ‘specified injury’, whilst this was 11.6% for equine clinical staff and 13.3% for production animal clinical staff. When considering their most severe injuries, the prevalence rose in all roles to 8.3% of non-clinical staff, 11.8% of mixed practice clinical staff, 25.4% of equine clinical staff and 20.0% of production animal clinical staff.

During their most recent injuries 50.0% of non-clinical staff were by themselves, which increased to 58.3% for their most severe injuries (Table 4). In contrast, for both recent and severe injuries 9% of clinical staff were working solo. For both recent and severe injuries 75% of non-clinical staff received medical treatment and 16.7% attended a hospital. In recent injuries, 68.3% of equine clinical staff received medical attention and 25.1% attended or were admitted to a hospital. This increased to 91.4% and 39.6% respectively for severe injuries. In recent injuries, two thirds of production animal clinical staff received medical attention and 40.0% attended or were admitted to a hospital. This increased to 85.7% and 57.2% respectively for severe injuries. In recent injuries, 55.6% of mixed practice clinical staff received medical attention and 5.6% attended a hospital. This increased to 88.8% and 58.9% respectively for severe injuries.

**Table 4.**
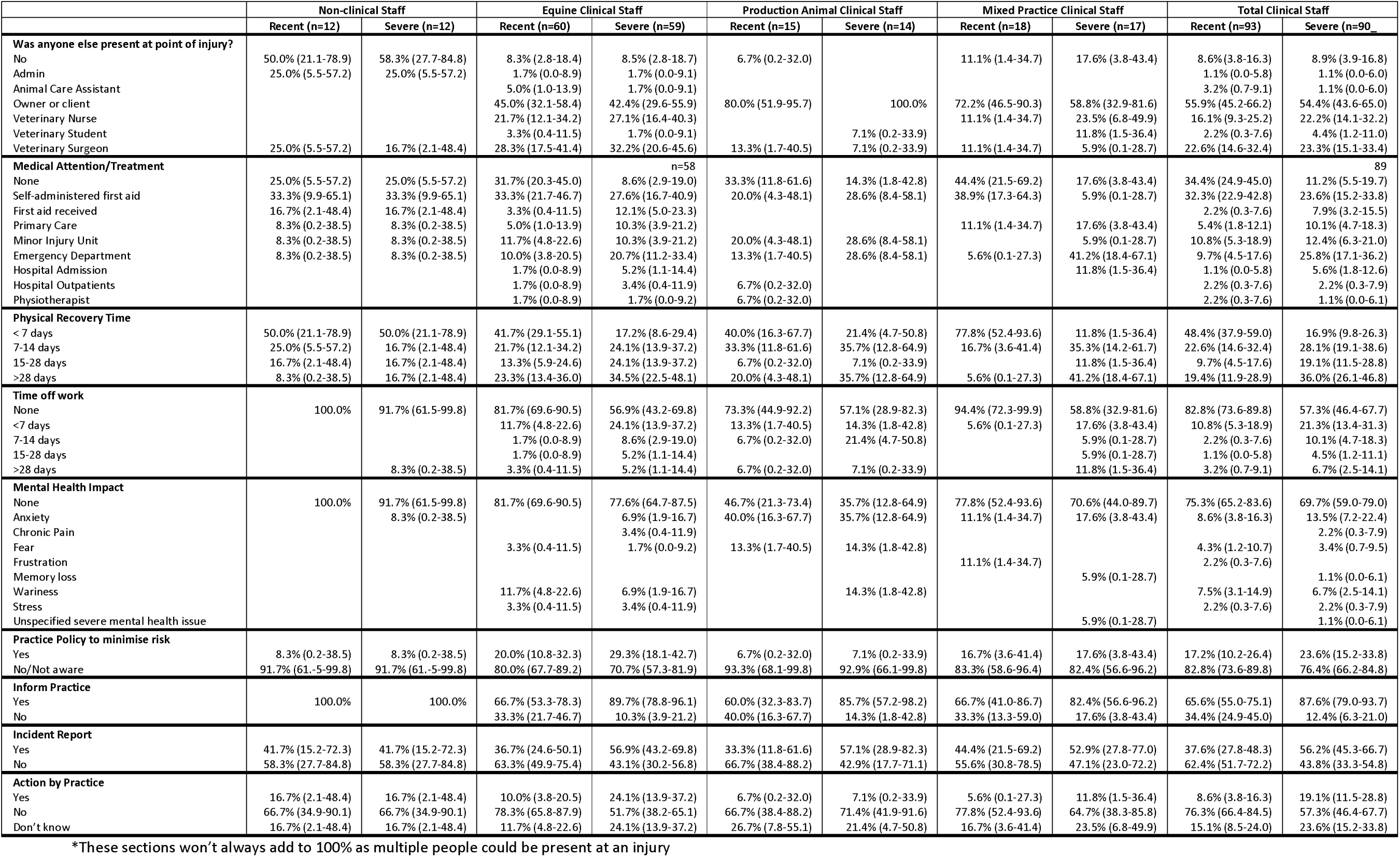
Consequences of work-related injuries in large animal practices.

In recent and severe injuries, 50.0% of non-clinical staff took more than seven days to physically recover from their injuries. However, in recent injuries none took any time off work, yet for a severe injury 8.3% took more than seven days off work, and therefore were reportable to HSE via RIDDOR. In recent injuries, 58.3% of equine clinical staff took more than seven days to physically recover, yet only 6.7% took more than seven days off work and were RIDDOR-reportable. For severe injuries, 82.8 had a longer recover, with 19.0% taking more than seven days off work. In recent injuries, 60.0% of production animal clinical staff took more than seven days to physically recover, yet only 13.3% took more than seven days off. For severe injuries, 78.6% had a longer recover, with 28.5% taking more than seven days off work. In recent injuries, 22.2% of mixed practice clinical staff took more than seven days to physically recover, yet none took more than seven days off work. For severe injuries, 88.2% had a longer recover, with 23.6% taking more than seven days off work. Most staff reported no mental health impacts from their injuries, except production animal clinicians where over half of injured persons reported a mental health impact. Across all roles, the most prevalent issues reported were anxiety and wariness of similar situations.

### Time off work

When asked why they did not take any time off work most non-clinical staff stated that their injury was not serious enough (S2 for content analysis tables and further supportive quotes) There were four themes emerging from equine clinical staff. The majority deemed that their injury was ‘not serious enough’. Many stated that their injury was ‘only a flesh wound’ (i.e. a serious injury that requires medical treatment and time off work, yet the injured person was dismissive of the seriousness of the situation) (Gilliam and Jones, 1975);

> *‘Cut a hole out of my trainer and carried on…’*

[Veterinarian, fractured foot resultant of a kick]

Some did not want to ‘let the team down’ and a few stated that they would ‘feel guilty’ about taking time off.

> *‘The show goes on. It wasn’t work limiting and didn’t want to put pressure on remaining staff’*

[Veterinarian, severely bruised leg resultant of a crush]

The below quote highlights the blasé tone that some individuals felt towards work-related injuries:

> *‘Lots of minor injuries happen within the equine world as it comes with working alongside dangerous half tonne animals. The type of person who works with such creatures know the risk & are hardy people for which a bite or being trodden on is a daily occurrence not phasing them.’*

Production animal clinical staff similarly said that they would not take time off work due to the injury not being serious enough, that it was ‘only a flesh wound’, and that they could still do restricted duties and didn’t want to let the team down.

> *‘It did hurt, and I could barely drive, but it was one of those things. You just continue working and think it will get better soon’*

[Veterinarian, severely bruised arm, cattle kick]

This cattle veterinarian who did take time off, due to a broken wrist resultant of a slip on a farm, voiced their concerns about their colleagues’ response to their injury:

> *‘Anxiety and fear that just would not regain full strength in my wrist. I was worried my boss would expect me to go back to full normal work as soon as my cast was removed and I would end up damaging my wrist further.’*

All mixed practice clinical staff stated that their injuries were not serious enough, however some additionally stated that they wanted to minimise their sick day usage, whilst others did not want to let the team down. This veterinarian received a shoulder strain from lifting a heavy animal and stated:

> *‘Although incredibly painful and I struggled to work - I do expect to get more injuries at work in the future due to the nature of being a mixed vet and working on farms with poor equipment, with horses, cattle, sheep, dogs, cats etc that are unpredictable or dangerous, driving so many miles each day etc. I know I am at increased risk of injury and I therefore want to minimise the amount of sick pay I use. We only get 10 days cumulative each year and I have already used 3 when I had flu recently.’*

### Change in behaviour

Most non-clinical staff would not their change behaviour if placed in a similar scenario preceding the injury (66.7% recent injuries, 75.0% severe injuries), but some would be more cautious when moving heavy objects (S3 for content analysis tables and further supportive quotes). Most clinical staff would change their behaviour, the exception being mixed practitioners when discussing their most recent injury. Examples of changes in behaviour for equine clinical staff included; adoption of protective headwear when working with horse, increased awareness of surroundings, and always sedating a horse when performing a procedure. Changes for production animal clinical staff included; being more cautious of crushes and handling equipment, changing how a procedure was performed, and being more aware of farm dogs. Changes for mixed practice clinical staff included; being more aware of surroundings, more aware of animal body language, generally being more cautious in the same scenario, and to always wear protective headwear when working with horses.

Some individuals reflected on the safety of working with animals without necessarily stating a specific behaviour change:

> *‘We changed the induction protocol at the time. I do not induce horses in a hospital setting anymore however when I anaesthetise horses I use a different technique and would now always wear a hard hat.’*

And a not inconsequential number of clinical staff would not change their behaviour following the injury they described.

> *‘Hard to prevent further injuries of this sort. The horse was well restrained and sedated’ ‘Have no idea how I managed to get knocked - always try to stay at side of front legs’*

> *‘I am more intolerant of junior colleagues taking silly risks with their welfare due to their ineptness with animal handling.’*

These quotes highlight a sense of inevitability, yet also surprise, that these injuries occurred.

### Practice policy, injury reporting, and actions by the practice

Most injured persons were not aware of any practice policy in place that would have minimised or removed the risk preceding the injury. When they did, they predominately mentioned a general health and safety policy, and equine clinical staff mentioned hard hat policies.

The majority of non-clinical staff (58.3% recent, 58.3% severe) and clinical staff report they (62.4% recent, 56.2% severe) did not fill in an incident report following their injury. The reasons for not filling in the reports highlighted that many of the injured parties do not understand the need to report work-related injuries. Reasons for not reporting included that it was too minor an injury to report, that they didn’t know they needed to, that it was a hazard of the job, they were too busy, and that reporting would make no difference.

> *‘I am old fashioned and have never reported an injury that didn’t require hospitalisation’*

[Veterinarian, back sprain, lifting a horse’s leg]

> *‘If I’m honest too much hassle for a known risk of the job and I’m not sure too much can be done’*

[Veterinarian, severe bruising leg, crushed by horse]

When asked whether there was any action taken by their veterinary practice after the injury, 16.7% of all staff stated there was a health and safety investigation. In some instances, this led to new working standard operating procedures and enforcement of PPE. One veterinary technician described the following action after being stood on by a horse:

> *‘I was encouraged several times by our clinical director and the health and safety advisor to contact the doctors/hospital as there was a query whether it was fractured-I declined’*

## Discussion

Work-related injuries are highly prevalent within the equine, production animal, and mixed practice veterinary workplaces, with many individuals being injured multiple times during their careers. Injuries were more prevalent in clinical staff, with most injuries occurring in a non-practice setting and were animal-related in nature. More than 1 in 5 injuries to clinical staff resulted in a hospital attendance, yet this was juxtaposed with minimal time off work to recover and little reporting of injuries to health and safety officers. This research highlights complex and often conflicting views towards managing safety and injuries in the veterinary workplace and is suggestive of a profession with a culture that does not prioritise health and wellbeing.

With over 90% of clinical staff working in this sector receiving a workplace injury during a career it is no wonder that some deem injury an inevitability. The lower exposure to non-clinical staff highlights that the nature of working with larger animals is at least partially responsible for the high injury rates. This is supported further by the result showing that those in clinical jobs are 36 time more likely to have received an injury than those in a non-clinical job. Interestingly, working out of hours was not associated with an increased odd of injury, however this maybe due to the analysis being underpowered as it was significant in univariable analysis. The annual prevalence is lower for equine veterinarians than anticipated, but equivalent for previous studies for production animal and mixed veterinarians (Mills, 2019). Age association with injury was more complex than anticipated. One would expect a linear relationship, as you could presume that the longer in a hazardous career the greater likelihood is that you have ever been injured, but the results show a non-linear relationship with a peak at 50-59 years old. This is hard to interpret and is likely due to many exposures and social factors, and may be influenced by how they personally define an injury (Furtado et al., 2024).

Those with non-clinical roles within this industry clearly highlight that these jobs are more clerical in nature with limited animal exposure. They are mainly minor injuries that occur within the practice premises and with injury mechanisms that are synonymous with office-based roles, such as slips and trips, manual handling, and being struck by office equipment (Health and Safety Executive, 2024b; NIOSH, 2024). As these injuries appeared to be genuinely minor, many had no treatment or solely received first aid.

The majority of situations (>80%) that resulted in an individual working in an equine clinical role being injured were involving a horse, unsurprisingly these were mainly in a location where the horse was being examined. Examinations or procedures involving the distal limb of the horse (36.7% of recent and 19.2% of severe injuries) were common events preceding an injury; similar to previous findings (Parkin et al., 2018). This is not surprising as in these situations an equine clinician is at their most vulnerable. They cannot see the horse’s face, which is the main organ that a horse uses to indicate that it is in discomfort or pain (van Loon and Van Dierendonck, 2018, 2017). A horse is likely to respond to pain by stamping, kicking, rearing or biting. Since in these examinations a veterinarian is effectively beneath the horse, any of these mechanisms could lead to severe injury. The consequences of being placed in vulnerable positions around the horse whilst performing an examination or procedure is highlighted by the common injury types (kicks, crushes and rears) (Parkin et al., 2018; Riley et al., 2015), and anatomy injured (leg, head, foot) (Eckert et al., 2011; Neville et al., 2024; Sritharan et al., 2022). The high proportion of head injuries and fractures are concerning, and potentially RIDDOR reportable injuries, especially when compared to the proportion of individuals that were wearing any form of PPE that may minimise the consequence of an injury (20% of recent injuries wore a helmet, 17% of severe). These head injuries are likely to be through impact with the environment (which is predominately concrete in a stable or yard) rather than through a direct kick to the head. Thus a helmet, would be protecting an individual from the impact of hitting these surfaces (Sritharan et al., 2022). Multiple studies have shown that head injuries are equally as likely when an individual is mounted or unmounted (Carmichael et al., 2014; Hoffmann et al., 2023). This, combined with the knowledge that those wearing helmets are at significantly lower risk of a severe brain injury when receiving head trauma than those that do not, mean that it is hard to justify the lack of helmet usage when working with horses (Bier et al., 2018; Carmichael et al., 2014).

The severity of many of these injuries is exemplified by the high hospitalisation rate; at least one in four injuries lead to hospital attendance. Thankfully, only a small proportion (<10%) were working solo at the point of injury, and so immediate first aid/responders is possible. These results highlight that veterinarians need ‘spotters’ to observe for a horse’s pain response and to warn veterinarians that a kick, stamp or rear could be about to occur. These ‘spotters’ will likely be a horse’s owner or stable hand, and training is therefore needed to establish the requirement for this role and educate on what signs they need to look for. Veterinarians themselves have called for ‘better handlers’ of their equine patients (Parkin et al., 2018). Additionally, helmet use should be standardised, there is no objective reason why helmets should not be worn whilst working with horses, and they greatly reduce injury consequences (Bier et al., 2018). The contrary health and safety behaviour of equine clinicians is illustrated by the time taken off-work being much lower than the time taken to physically recover. This is similar to production animal and mixed clinicians and is discussed below.

Production animal clinicians were predominately injured by cattle whilst working on farm, though there were a surprising number of dog bite injuries. The low number of respondents made contextual insight challenging and further research is needed. Other studies have identified time spent in close proximity to cattle increases the chances of injury (Lindahl et al., 2013; “The Effect of Stress, Attitudes, and Behavior on Safety during Animal Handling in Swedish Dairy Farming,” 2015), something which is hard to avoid as a veterinarian. Contextually, clinical examinations and veterinary procedures have been previously identified as higher risk situations (Tulloch et al., 2023; Wilkins et al., 2009) The injuries sustained were significant with many fractures and crush type injuries. The hospitalisation rate was very high, similar to other agricultural professions injured by cattle (Ehrhard et al., 2022; Fritschi et al., 2006; Lucas et al., 2013; Murphy et al., 2010; Rhind et al., 2021), with resultant recovery periods being long. This is a reflection on the seriousness of the injuries, yet little time was taken off work. Production animal clinicians were the only group to highlight significant mental health impacts, namely anxiety and wariness of similar situation. It is unknown why this would differ from other types of clinicians, especially since one of the only studies exploring mental health in different veterinary sectors found that production animal veterinarians had the lowest prevalence of severe mental health issues (Nett et al., 2015). At no point did this extensive survey identify that injury and illness impacts the mental health of veterinarians. Our novel finding necessitates further research. Mixed clinicians were injured in more varied locations, with more species involved, however the farm was the most common location. In recent injuries few reported having medical treatment, however more than half attended hospital for their severe injuries. These injuries had lengthy recovery periods, with few taking time off work.

Collectively this sector of the veterinary industry would change their behaviour if placed in the same scenario in the future, often through the usage of PPE, being more aware of surroundings or using better restraint (either pharmacological or physical). However, a concerning proportion would not change their behaviours and would enter the same situation in the same way. There was a strong sense of inevitability of these events happening and that by joining this profession these injuries were inescapable. This is indicative of a lack of reflection and rather blasé attitude towards injuries, as well as potentially an overconfidence of their handling skills (Davies et al., 2022; Titterington et al., 2022). There is the potential that, like musicians, these attitudes are driven by seeing injury as a sign of weakness and failure, and so the non-disclosure of injuries and lack of change in behaviour become commonplace (Rickert et al., 2014). Additionally, being open about injuries could lead to judgement from their peers. It is indicative that there is a lack of reflection, with no desire to improve and learn from the injuries that they have received. This is a deep cultural issue, that requires further research to understand why this persists, and lessons need to be learnt from other industries who have successfully changed their injury culture (Cooper Ph.D., 2000; Duryan et al., 2020; Nielsen, 2014; Zuschlag et al., 2016)

Many individuals did not take time off work despite serious injuries that required more than a week to physically recover from. This minimisation of the injury captured by the phrase ‘only a flesh wound’ is interlinked with feelings of guilt and not wanting to ‘let the team down’. This culture of presenteeism starts at veterinary schools (Furtado et al., 2024; Kinman and Clements, 2023; Riley et al., 2015; Tulloch et al., 2023; Whittem et al., 2021), and appears to be widespread across the profession, this is exemplified by veterinarians stating that something is only an injury if it means they cannot do their job properly (Furtado et al., 2024). This is also present within the agricultural industry, where associated risk factors include; poor work safety climate, higher percentage of female workers, and worker dissatisfied with management (Siqueira et al., 2023). Within the equestrian sector many do not accept the risk of the job, injuries are normalised and ignored, and only reported and discussed if severe, with high levels of presenteeism (Davies et al., 2022; Lindahl et al., 2022). Presenteeism was explained by financial circumstances, perceived staff shortages, previous injury experiences, and perceived employer expectations (Davies et al., 2022), similar to what we might expect for equine veterinarians. It was concerning that limited sick days made some individuals not report their injury nor take time off work. The workplace culture needs to shift to empower employees to discuss this with their employer and develop an appropriate time off work and return to work strategy. It will be challenging to reduce presenteeism unless organisational culture prioritises employee health and workplace injuries are not stigmatised (Kinman and Clements, 2023). This can be leveraged through, ensuring employees are not over-worked, managers asking people to take time off work, leaders not working when injured, and managing expectations of workloads on return to work.

The RIDDOR legislation and reporting is the main way of reporting injuries in the UK and generating national injury figures, this is critical for relatively small professions, such as the veterinary industry. Yet the discordance in time to recovery and time off work, will mean that injuries that are severe enough for an individual to take more than a week off work are not captured as the injured party won’t take time off for recovery. Additionally, the self-reported low levels of injury reporting will mean that even ‘specified injuries’ will not be reported. Further, like agricultural workers, incidents are less likely to be reported if the individual involved does not want to admit irresponsible working or feel shamed by what occurred (Titterington et al., 2022). It is likely that the UK has very inaccurate figures regarding veterinary workplace injuries, in particular within the settings studied in this research, as they just aren’t being captured. This can be seen in the RIDDOR national data where the incidence of veterinary ‘specified injuries in 2022-23 was 97 injuries per 100,000 employees (the national average was 63, and the top incidence was 451), and the incidence of over-7-day injury per 100,000 employees was 196 (the national average of 152, and top of 1336) (Health and Safety Executive, 2024b). Our data supports that RIDDOR reported events are highly under-reported. To simplify this, if an equine or livestock veterinarian gets injured, and their ability to work is not impeded or it doesn’t lead to a hospital attendance, they won’t consider it to be an injury, they then will not take time off work to recover nor report it to their employer. Thus, continuing to work while injured, putting themselves and others at further risk. Subsequently, accurate injury statistics are not generated, making the industry seem safer than it is, resulting in no indicators that improvement and change to the safety culture is needed, and just further empowering the status quo.

The low response rate and resultant sample size are a limitation of this study. Combining this with respondents being solely from one consolidated group of veterinary practices could raise questions about the representativeness of the study population. The respondent demographics are broadly representative of the veterinary clinical population and the nature of injurie described are similar to previous research and so we believe the results are indicative of what is occurring within the veterinary industry. However, to further validate these results more wide-spread surveys and/or interviews and focus groups are needed. There may be two forms of response bias present. Firstly, that individuals with a history of a work-related injury may have been more likely to respond, potentially elevating disease prevalence figures. We attempted to alleviate this by incentivising all responses. Secondly, the survey was disseminated via the participants employee and their answers may reflect what they thought their employer wanted to hear. We aimed to minimise this by reminding respondents that no responses would be seen or analysed by their employer and all responses were anonymous.

The non-centralised structure of the veterinary industry means that a systematic approach to improve safety culture and reduce the number of incidents, and their severity is going to be challenging. A multifaceted approach that improves attitudes, behaviours, systems, and practices is needed, and this will need significant support and investment (Cooper Ph.D., 2000; Fitzgerald, 2005; Nielsen, 2014). To create a safety mindset the industry needs to see safety as a core value that is embedded in organisational culture; leadership is key to this. Leaders must prioritise safety, for example having all meetings starting with a safety review, provide sufficient resources for safety initiatives, promote positive reinforcement, and highlight that safety is a shared responsibility (Cooper Ph.D., 2000; Nielsen, 2014; Zuschlag et al., 2016). Agricultural coalitions have shown that effective leadership improves the safety culture on farms (Titterington et al., 2022; Wendl and Cramer, 2018). To succeed, effective communication is vital. There needs to be open communication where accountability is encouraged, blame is minimised, and with no fear of retribution. Employees should be empowered to take responsibility for their own safety and for the safety of their colleagues and clients. There needs to be mechanisms were effective and actionable feedback can be raised and delivered (Nielsen, 2014; Zuschlag et al., 2016). These communication strategies must be appropriate for the local context and can lead to improved general health, less depression, higher job and life satisfaction, and less productivity loss of employees (Katz et al., 2019).

Reporting systems for injuries and near-misses are integral but will only succeed if the need for reporting is understood by the industry (Fitzgerald, 2005; Titterington et al., 2022). Regular audits and inspections should be performed, with continuous improvement prioritised. To support this safety performance metrics, need to be developed. Campaigns to promote awareness and encourage the need of reporting need to be considered, especially since the industry has such low reporting rates (Zuschlag et al., 2016). Comprehensive training around safety protocols, risk management and first aid/responder training, which are regularly updated, and utilises scenario-based learning should be developed and utilised to raise awareness and to build baseline competencies (Duryan et al., 2020). Employees should be involvement with the development and review of safety policies and practices, to ensure all the views and scenarios of all those in the industry are considered (Titterington et al., 2022). Care should be taken as some may value personal experience more than education and as such may not engage with training. Safety recognition programmes should be introduced that reward individuals or practices who demonstrate exceptional commitment to safety practices. All injuries and near-misses should be investigated with lessons learned shared across organisations to prevent recurrence. Finally, technology and personal protective equipment should be utilised when appropriate. There is no acceptable reason why hard hats cannot be worn when working around horses, this will minimise the consequence of any head trauma and will show the wider equine community that safety is taken seriously within the veterinary profession (Carmichael et al., 2014; Neville et al., 2024). Many practices are well on the way in developing these cultures and systems, however, to have the widespread change that is needed is going to be a big task. Attitudes to injury are outdated and unsafe, and modernisation is urgently needed.

## Conclusions

There is no doubt that veterinarians who work with horses and livestock have a dangerous job, but potentially their attitudes are more dangerous. Work-related injuries are alarmingly common within this sector with clinical staff frequently injured, often severely. Most injuries occur outside the practice, are animal-related, and often lead to hospital visits, yet are rarely reported, reflecting a work culture that undervalues health and safety. This culture normalises injury as inevitable. The lack of protective measures, such as helmet use, and the tendency to continue working despite serious injuries, highlight a need for urgent cultural and systemic change. Improved safety practices, leadership commitment, and comprehensive training are essential to address this deep-seated issue and reduce the high injury rates in large animal veterinary workplaces.

## Data Availability

All data produced in the present study are available upon reasonable request to the authors

## Acknowledgements

The authors would like to thank all the participants of this study for their frank and honest viewpoints.

## Author Contributions

Conceptualisation: JT. Funding Acquisition – JT, MW, RJ. Methodology: JT, MW, IS. Formal analysis: JT. Writing original draft – JT. Writing review & editing – All authors

---

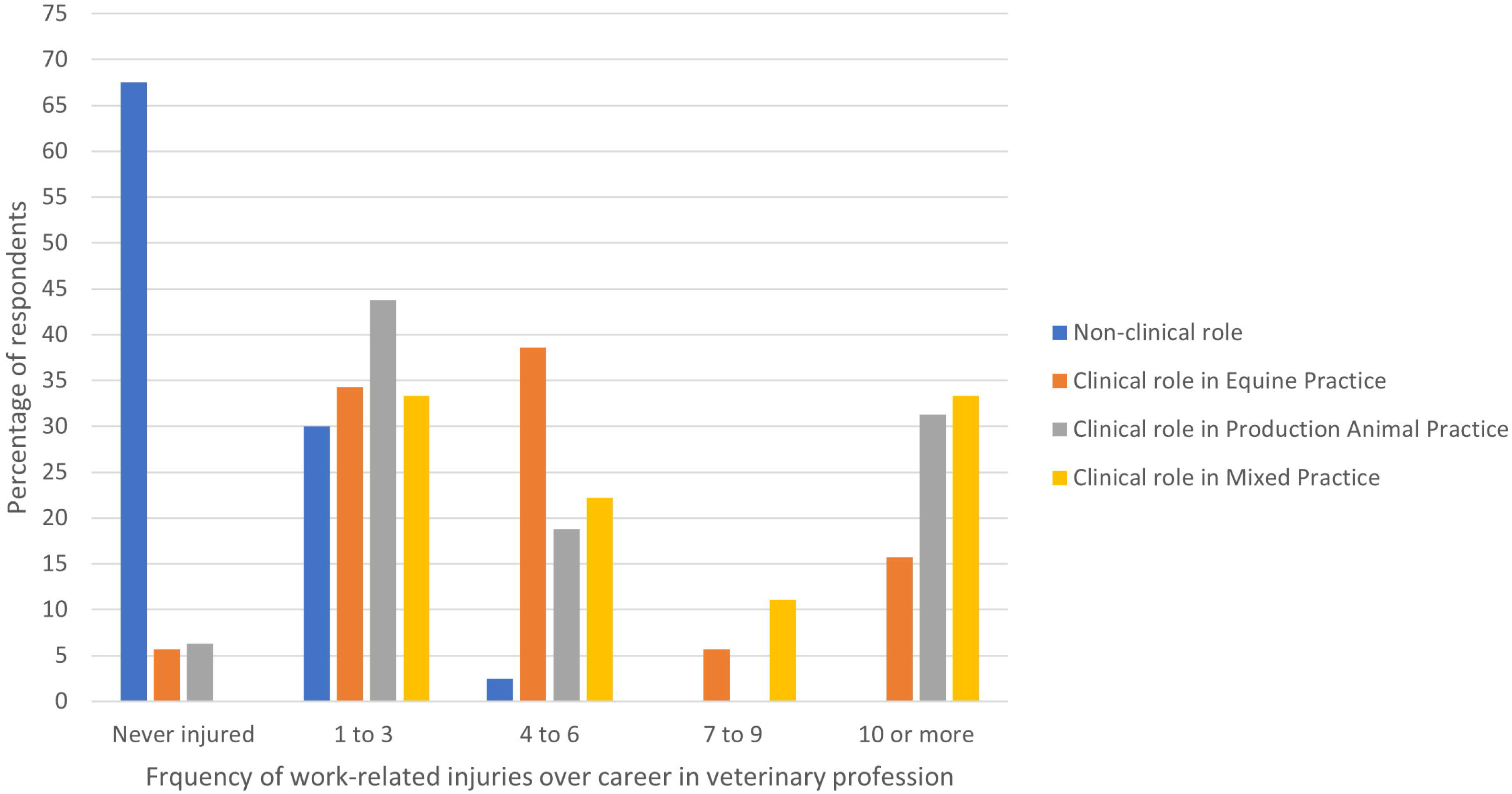

