## Supplementary Material for "‘It’s only a flesh wound’ – understanding the safety culture in equine, production animal and mixed veterinary practices"

S1 - Mechanism and resultant injury of animal-related veterinary work-related injuries.

| Mechanism | Body part | Injury | Equine Clinical Staff | | Farm Clinical Staff (n=12) | | Mixed Practice Clinical Staff (n=16) | |
|---|---|---|---|---|---|---|---|---|
| | | | **Recent (n=50)** | **Severe (n=50)** | **Recent (n=12)** | **Severe (n=8)** | **Recent (n=16)** | **Severe (n=9)** |
| **Bite** | | | **4.0% (0.5-13.7)** | **6.0% (1.3-16.6)** | **25.0% (5.5-57.2)** | **25.0% (3.2-65.1)** | **37.5% (16.8-62.4)** | **55.6% (21.2-86.3)** |
| | *Arm* | | | | | | | |
| | | Bruising | | | | | 6.3% (0.2-30.2) | 33.3% (7.5-70.1) |
| | | Puncture | | | 8.3% (0.2-38.5) | 12.5% (0.3-52.7) | | |
| | *Hand* | | | | | | | |
| | | Laceration | 4.0% (0.5-13.7) | 4.0% (0.5-13.7) | | 12.5% (0.3-52.7) | 6.3% (0.2-30.2) | |
| | | Puncture | | | 16.7% (2.1-48.4) | | 25.0% (7.3-52.4) | 22.2% (2.8-60.0) |
| | *Multiple* | | | | | | | |
| | | Bruising | | 2.0% (0.1-10.7) | | | | |
| **Burn (Rope)** | | | | | | **12.5% (0.3-52.7)** | | |
| | *Hand* | | | | | | | |
| | | Fracture | | | | 12.5% (0.3-52.7) | | |
| **Crush** | | | **22.0% (11.5-36.0)** | **12.0% (4.5-24.3)** | **41.7% (15.2-72.3)** | **25.0% (3.2-65.1)** | **18.8% (4.0-45.7)** | **22.2% (2.8-60.0)** |
| | *Arm* | | | | | | | |
| | | Sprain | 2.0% (0.1-1.07) | 2.0% (0.1-10.7) | | | | |
| | *Chest* | | | | | | | |
| | | Bruising | 2.0% (0.1-1.07) | | | | | |
| | | Fracture | 2.0% (0.1-1.07) | 4.0% (0.5-13.7) | | | | |
| | *Foot* | | | | | | | |
| | | Bruising | 6.0% (1.3-16.6) | | 16.7% (2.1-48.4) | | 12.5% (1.6-38.4) | |
| | | Fracture | 2.0% (0.1-1.07) | | | 12.5% (0.3-52.7) | | 11.1% (0.3-48.3) |
| | *Hand* | | | | | | | |
| | | Bruising | 2.0% (0.1-1.07) | | 8.3% (0.2-38.5) | | 6.3% (0.2-30.2) | |
| | | Fracture | | | 8.3% (0.2-38.5) | 12.5% (0.3-52.7) | | 11.1% (0.3-48.3) |
| | | Laceration | | | 8.3% (0.2-38.5) | | | |
| | *Head* | | | | | | | |
| | | Concussion | | 2.0% (0.1-10.7) | | | | |
| | *Leg* | | | | | | | |
| | | Bruising | 2.0% (0.1-1.07) | | | | | |
| | | Fracture | 2.0% (0.1-1.07) | 2.0% (0.1-10.7) | | | | |
| | | Sprain | 2.0% (0.1-1.07) | | | | | |
| | *Pelvis* | | | | | | | |
| | | Bruising | | 2.0% (0.1-10.7) | | | | |
| **Ergonomic** | | | **8.0% (2.2-19.2)** | **2.0% (0.1-10.7)** | **8.3% (0.2-38.5)** | | **12.5% (1.6-38.4)** | |
| | *Back* | | | | | | | |
| | | Sprain | 4.0% (0.5-13.7) | 2.0% (0.1-10.7) | 8.3% (0.2-38.5) | | 6.3% (0.2-30.2) | |
| | *Hand* | | | | | | | |
| | | Fracture | 2.0% (0.1-1.07) | | | | | |
| | *Shoulder* | | | | | | | |
| | | Sprain | 2.0% (0.1-1.07) | | | | 6.3% (0.2-30.2) | |
| **Fall** | | | | **2.0% (0.1-1.07)** | | | | |
| | *Pelvis* | | | | | | | |
| | | Bruising | | 2.0% (0.1-1.07 | | | | |
| **Headbutt** | | | **4.0% (0.5-13.7)** | **4.0% (0.5-13.7)** | | | | |
| | *Head* | | | | | | | |
| | | Bruising | 2.0% (0.1-1.07) | 2.0% (0.1-1.07) | | | | |
| | | Concussion | 2.0% (0.1-1.07) | 2.0% (0.1-1.07) | | | | |
| **Kick** | | | **56.0% (41.3-70.0)** | **56.0% (41.3-70.0)** | **25.0% (5.5-57.2)** | **37.5% (8.5-75.5)** | **31.3% (11.0-58.7)** | **22.2% (2.8-60.0)** |
| | *Abdomen* | | | | | | | |
| | | Bruising | | 2.0% (0.1-10.7) | | 12.5% (0.3-52.7) | | |
| | *Arm* | | | | | | | |
| | | Bruising | 2.0% (0.1-1.07) | 2.0% (0.1-10.7) | | | | |
| | | Fracture | | 2.0% (0.1-10.7) | | | | |
| | *Chest* | | | | | | | |
| | | Fracture | 2.0% (0.1-1.07) | | | | | |
| | *Foot* | | | | | | | |
| | | Bruising | 2.0% (0.1-1.07) | | | | | |
| | | Fracture | 2.0% (0.1-1.07) | | | | | |
| | *Hand* | | | | | | | |
| | | Fracture | 2.0% (0.1-1.07) | 2.0% (0.1-10.7) | | | | |
| | | Sprain | 2.0% (0.1-1.07) | | | | | |
| | *Head* | | | | | | | |
| | | Bruising | 2.0% (0.1-1.07) | 2.0% (0.1-10.7) | | | 12.5% (1.6-38.4) | |
| | | Concussion | 6.0% (1.3-16.6) | 10.0% (3.3-21.8) | | | | |
| | | Fracture | 2.0% (0.1-1.07) | 2.0% (0.1-10.7) | | | | |

| | | | | | | | | |
|---|---|---|---|---|---|---|---|---|
| | | Laceration | 2.0% (0.1-1.07) | 2.0% (0.1-10.7) | | | | |
| | *Leg* | Bruising | 24.0% (13.1-38.2) | 16.0% (7.2-29.1) | 8.3% (0.2-38.5) | 12.5% (0.3-52.7) | 18.8% (4.0-45.7) | 22.2% (2.8-60.0) |
| | | Fracture | | 2.0% (0.1-10.7) | 8.3% (0.2-38.5) | | | |
| | | Laceration | 6.0% (1.3-16.6) | | | | | |
| | | Sprain | | 2.0% (0.1-10.7) | | | | |
| | *Multiple* | Bruising | 2.0% (0.1-1.07) | 2.0% (0.1-10.7) | | | | |
| | | Concussion | | | 8.3% (0.2-38.5) | 12.5% (0.3-52.7) | | |
| | | Fracture | | 2.0% (0.1-10.7) | | | | |
| **Knocked over** | | | **4.0% (0.5-13.7)** | | | | | |
| | *Leg* | Bruising | 2.0% (0.1-1.07) | | | | | |
| | *Pelvis* | Bruising | 2.0% (0.1-1.07) | | | | | |
| **Rear** | | | | **18.0% (8.6-31.4)** | | | | |
| | *Back* | Disc Protrusion | | 2.0% (0.1-10.7) | | | | |
| | *Hand* | Fracture | | 2.0% (0.1-10.7) | | | | |
| | | Sprain | | 2.0% (0.1-10.7) | | | | |
| | *Foot* | Bruising | | 4.0% (0.5-13.7) | | | | |
| | | Fracture | | 4.0% (0.5-13.7) | | | | |
| | *Shoulder* | Fracture | | 2.0% (0.1-10.7) | | | | |
| | *Multiple* | Sprain | | 2.0% (0.1-10.7) | | | | |
| **Scratch** | | | **2.0% (0.1-1.07)** | | | | | |
| | *Hand* | Laceration | 2.0% (0.1-1.07) | | | | | |

### S2 – Time off Work Themes and supportive quotes

| | **Non-clinical Staff** | | **Equine Clinical Staff** | | **Production animal Clinical Staff** | | **Mixed animal Clinical Staff** | |
|---|---|---|---|---|---|---|---|---|
| | **Recent** | **Severe** | **Recent** | **Severe** | **Recent** | **Severe** | **Recent** | **Severe** |
| 'Not serious enough' | 83.3% | 81.8% | 63.3% | 54.5% | 36.3% | 37.5% | 100% | 100% |
| 'Only a flesh wound' | | | 28.6% | 18.2% | 36.3% | 25.0% | | |
| 'Don't want to let the team down' | | | 10.2% | 15.2% | 27.3% | 25.0% | | 28.6% |
| Feelings of guilt about taking time off work | | | 0% | 12.1% | | | | |
| Minimise sick day usage | | | | | | | 14.3% | |

#### 'Only a flesh wound'

*'Just got on with it. Wasn't really sore until the next day so carried on'*

[Equine Veterinary nurse, widespread bruising due to being reared on]

'Although it was very painful at the time it did not prevent me from driving or walking and healed within a few days'

[Production Animal Veterinarian, bruised foot, cattle crush]

**‘Don’t want to let the team down’**

*‘Would have meant additional strain on the remaining team. I was just about able to drive so could work’*

[Production Animal Veterinarian, bruised leg resultant of a kick]

*‘It did not affect my ability to perform my job and the practice was incredibly busy so I did not want to overload my colleagues by taking time off work unnecessarily.’*

[Livestock Veterinarian, hand laceration, dog bite]

**Minimise sick day usage/ Feelings of guilt about taking time off work**

*‘Concerns about sick pay during recovery period as unable to work, stress/pressure to return to work, no option for desk duties. Frustration, depression, annoyance, chronic arthritis. Upset during painful flare ups. Concern for long-term working capacity as struggle with dexterity and strength - have adapted for surgical procedures but still struggle.’*

[Mixed-animal Veterinarian, fractured hand, equine procedure]

**S3 – Change in behaviour themes**

| | Non-clinical Staff | | Equine Clinical Staff | | Production animal Clinical Staff | | Mixed animal Clinical Staff | |
|---|---|---|---|---|---|---|---|---|
| | Recent | Severe | Recent | Severe | Recent | Severe | Recent | Severe |
| No change | 66.7% | 75.0% | 38.3 | 24.1 | 40.0% | 28.6% | 55.6% | 29.4 |
| Caution moving heavy objects | 25.0% | 25.0% | | | | | | |
| Adoption of protective headwear | | | 20.0 | 20.7 | | | | 11.8% |
| Increased awareness of surroundings | | | 13.3% | 13.8% | | | | 17.6% |
| Always sedating a horse when undertaking a procedure | | | | 10.3% | | | | |
| More cautious of crushes and handling equipment | | | | | 33.3% | 21.4% | | |
| Change how a procedure was performed | | | | | | 21.4% | | |

| | | | | | | | | |
|---|---|---|---|---|---|---|---|---|
| More aware of farm dogs | | | | | 13.3% | | | |
| More aware of animals body language | | | | | | | | 11.8% |
| More cautious in same scenario | | | | | | | 11.1 | 11.8 |

**Further Quotes:**

**Self-prompted reflections on safety**

*‘‘As I get older this sort of occurrence does make me question the safety working with horses’*

*‘I am very aware of the damage that can be caused by horse bites and I am now especially careful. In addition I tell all students and interns that I train to pay attention to the front end and that animals in pain react differently.’*

*‘Refuse to put my hands/body in the line of fire when cattle are wild or improperly restrained’*

**Reasons for no change in behaviour**

*‘No - accidental injury due to the unpredictability of animals’*

*‘No - I would not put on any protective equipment’*

*‘More cautious with headcollars and cattle, but still also don't want to put them in the crush because they sit down and get stuck. But if cow dangerous less hesitant to take the risk and put them in the crush now’*
